# Development and Characterization of Proteomic Aging Clocks in the Atherosclerosis Risk in Communities (ARIC) Study

**DOI:** 10.1101/2023.09.06.23295174

**Authors:** Authors: Shuo Wang, Zexi Rao, Rui Cao, Anne H. Blaes, Josef Coresh, Corinne E. Joshu, Benoit Lehallier, Pamela L. Lutsey, James S. Pankow, Sanaz Sedaghat, Weihong Tang, Bharat Thyagarajan, Keenan A. Walker, Peter Ganz, Elizabeth A. Platz, Weihua Guan, Anna Prizment

## Abstract

Biological age may be estimated by proteomic aging clocks (PACs). Previous published PACs were constructed either in smaller studies or mainly in White individuals, and they used proteomic measures from only one-time point. In the Atherosclerosis Risk in Communities (ARIC) study of about 12,000 persons followed for 30 years (around 75% White, 25% Black), we created de novo PACs and compared their performance to published PACs at two different time points. We measured 4,712 plasma proteins by SomaScan in 11,761 midlife participants, aged 46-70 years (1990-92), and 5,183 late-life pariticpants, aged 66-90 years (2011-13). All proteins were log2-transformed to correct for skewness. We created de novo PACs by training them against chronological age using elastic net regression in two-thirds of healthy participants in midlife and late life and compared their performance to three published PACs. We estimated age acceleration (by regressing each PAC on chronological age) and its change from midlife to late life. We examined their associations with mortality from all-cause, cardiovascular disease (CVD), cancer, and lower respiratory disease (LRD) using Cox proportional hazards regression in all remaining participants irrespective of health. The model was adjusted for chronological age, smoking, body mass index (BMI), and other confounders. The ARIC PACs had a slightly stronger correlation with chronological age than published PACs in healthy participants at each time point. Associations with mortality were similar for the ARIC and published PACs. For late-life and midlife age acceleration for the ARIC PACs, respectively, hazard ratios (HRs) per one standard deviation were 1.65 and 1.38 (both p<0.001) for all-cause mortality, 1.37 and 1.20 (both p<0.001) for CVD mortality, 1.21 (p=0.03) and 1.04 (p=0.19) for cancer mortality, and 1.46 and 1.68 (both p<0.001) for LRD mortality. For the change in age acceleration, HRs for all-cause, CVD, and LRD mortality were comparable to those observed for late-life age acceleration. The association between the change in age acceleration and cancer mortality was insignificant. In this prospective study, the ARIC and published PACs were similarly associated with an increased risk of mortality and advanced testing in relation to various age-related conditions in future studies is suggested.

## Introduction

In the United States the average human life expectancy has increased by 30 years during the 20^th^ century. This increased life expectancy has given rise to the number of individuals living with age-related diseases and disabilities and has inevitably led to an increased risk of mortality, reduced health span, lower quality of life, and increased healthcare costs in the United States. Research is needed to understand the biological mechanisms of aging as we develop and target preventions and interventions that prolong healthy lifespan (1, 2).

An individual’s extent of aging, i.e., how far they are into the aging process, cannot be sufficiently measured by chronological age as individuals develop physiological dysregulations at different chronological ages (1, 2). To better understand the extent of aging, researchers introduced a term called “biological age” to capture how far individuals are into their aging process independent of chronological age. Biological age, according to the definition proposed by Baker and Sprott, is characterized by the “biological parameter[s] of an organism, either alone or in some multivariate composite that will, in the absence of disease, better predict functional capability at some late age than will chronological age” (3).

To estimate a person’s biological age, researchers have developed metrics called aging clocks using epigenetic, transcriptomic, metabolomic, proteomic, and other biomarkers (4). Aging clocks are strongly correlated with chronological age in healthy individuals. However, in individuals with comorbidities or predisposing conditions, aging clocks deviate from chronological age because these conditions impact levels of age-associated biomarkers (5, 6). Studies show that aging clocks may be used to identify individuals who have a positive deviation of biological age from their chronological age (called age acceleration) that may predict their future risk of age-related conditions (5–7). In addition, aging clocks may also track the effectiveness of anti-aging interventions in clinical trials (5, 8–10).

The most studied aging clocks are epigenetic clocks, such as the Horvath clock, Hannum clock, DNAm PhenoAge, and GrimAge (11–14). However, there is a lack of understanding of the underlying mechanisms of aging-related changes in DNA methylation sites. It remains unclear what aspects of aging those clocks reflect (15). Recently, new assays that measure thousands of proteins in a small blood sample simultaneously have been developed. For instance, the SomaScan assay, a modified aptamer-based technology (16–18). These assays make it possible to construct proteomic aging clocks (PACs) (5–7, 19). The strength of PACs is that they include proteomic-based biomarkers, an intermediate phenotype that is most proximal to age-related diseases, and thus may provide more accurate information on aging and age-related pathologies (5, 20). Importantly, proteins serve as a target in 96% of FDA-approved drugs (21). Therefore, in addition to predicting biological age and risk of diseases, proteins comprising PACs, if causal, hold promise as targets of anti-aging drugs. Targeting age-related processes or pathological manifestations instead of a single disease is advantageous as this approach may simultaneously reduce the development or progression of multiple age-related diseases and potentially prolong healthy lifespan.

Several PACs have been developed using SomaScan assays, such as the PACs created by Lehallier [2020] (N = 3,301, aged 18-76 years) (6), Tanaka [2018] (N = 240, aged 22-93 years) (5), and Sathyan [2020] (N = 1,025, aged 65-95 years) (19). The descriptions of those published PACs, including the number of proteins used to construct those PACs, are presented in **S1 Table**. Although those published PACs showed high correlations with chronological age, they were developed either in relatively small studies or in studies included individuals of European descent (5, 6, 19, 22). However, proteins associated with age and age-related diseases vary by race and socioeconomic status (23–25). Moreover, previously published PACs were constructed using a one-time measure. Thus, it is necessary to develop PACs in a large longitudinal study of diverse individuals and examine if the change in PACs over time is associated with mortality independent of chronological age, smoking, and other lifestyle factors and behaviors.

In this study, we developed new PACs in participants followed from midlife and late life and examined their associations with mortality within a large population-based prospective cohort of White and Black, men and women, in the Atherosclerosis Risk in Communities (ARIC) study. In ARIC, about 5,000 plasma proteins were measured using the SomaScan assay (v.4) from plasma samples collected at two different times (20 years apart). We aimed to compare the midlife and late-life ARIC PACs developed in healthy participants (without major age-associated diseases) with the published Lehallier’s, Tanaka’s, and Sathyan’s PACs. Comparing their correlation with chronological age and their associations with mortality from all-cause, cardiovascular disease (CVD), cancer, and lower respiratory disease (LRD). In addition, using protein data measured at two different time points, we examined whether the change in PACs from midlife to late life was associated with premature mortality.

## Methods

### Study population

This study included White and Black, men and women, participants of the ongoing ARIC study (RRID: SCR_021769), which was initiated in 1987 (26, 27). At Visit 1 (1987–89), 15,792 volunteers aged 45-64 years were recruited from four U.S. study centers, Washington County, Maryland; the northwest suburbs of Minneapolis, Minnesota; Jackson, Mississippi; and Forsyth County, North Carolina. Participants in the Minnesota and Maryland centers were primarily White and the recruitment in Mississippi was restricted to Black residents. ARIC was approved by institutional review boards at each participating center and all study participants provided written informed consent. To date, nine visits have been completed (26). ARIC participants have received follow-up telephone calls annually from 1987 to 2012 and semi-annually after 2012, with response rates of 90%-99% for the annual follow-up calls and 83%-90% for semi-annual follow-up calls among living participants who have not withdrawn consent to be contacted (27). There is also continuous surveillance of local hospitals and linkage to the National Death Index (NDI).

### Plasma collection

In this study, we used plasma samples collected at Visit 2 (1990–92) from 11,761 participants aged 46-70 years (midlife) and at Visit 5 (2011–13) from 5,183 participants aged 66-90 years (late life). The blood sample collection, processing, and storage in ARIC was designed to minimize the spontaneous biochemical reactions after blood collection and is consistent with the recommended practice for proteomics data analysis in epidemiological studies (16, 28, 29). After venipuncture, blood samples were put immediately in an ice water bath. Centrifugation was performed within 10 min after venipuncture at room temperature (15-25 °C). After centrifugation, the aliquots were stored at −80 °C within 90 min from venipuncture and were unthawed before this analysis.

### Protein measurement and quality control

Plasma samples were analyzed using a SOMAmer (Slow Off-rate Modified Aptamers) based capture array called SomaScan® by Somalogic, Inc. (Boulder, CO, USA) (18, 30–32). The SomaScan platform uses single-stranded modified DNA-based aptamers to capture conformational protein epitopes. The description of the SomaScan assay and the data normalization process have been described previously (16, 17, 32).

Among the 5,284 available aptamers, we excluded aptamers with a Bland-Altman coefficient of variation (CVBA) greater than 50% or a variance of less than 0.01 on the log scale, or binding to mouse Fc-fusion, contaminants, or non-proteins (33). After the exclusion, 4,955 aptamers were included (at Visit 2 and Visit 5) which corresponded to 4,712 proteins. About 5% of proteins had more than one aptamer binding to the same protein. Each aptamer was treated as a variable in the construction of PACs. The CVBA for split samples was 6% at Visit 2 and 7% at Visit 5. Protein measures were reported as relative fluorescent units (RFU) and were log2-transformed to correct for skewness.

### Identifying healthy participants

In this study, we created the midlife (Visit 2) and late-life (Visit 5) ARIC PACs in “healthy participants” defined as participants without major age-associated diseases that are linked to premature mortality. Specifically, abnormal kidney function (i.e., estimated glomerular filtration rate (eGFR) less than 60 mL/min/1.73m^2^), cancer, chronic obstructive pulmonary disease (COPD), CVD (heart failure, definite or probable stroke, or coronary heart disease (34, 35)), diabetes, and hypertension (or uncontrolled hypertension for late-life participants at Visit 5). The definitions and assessments of these major diseases in ARIC and the detailed process of identifying healthy participants are described in the **S1 Appendix**. We identified 4,489 midlife healthy participants at Visit 2 (38.2% of all Visit 2 participants, **Fig 1**) and 945 late-life healthy participants at Visit 5 (18.2% of all Visit 5 participants, **Fig 2**).

**Figure 1.**
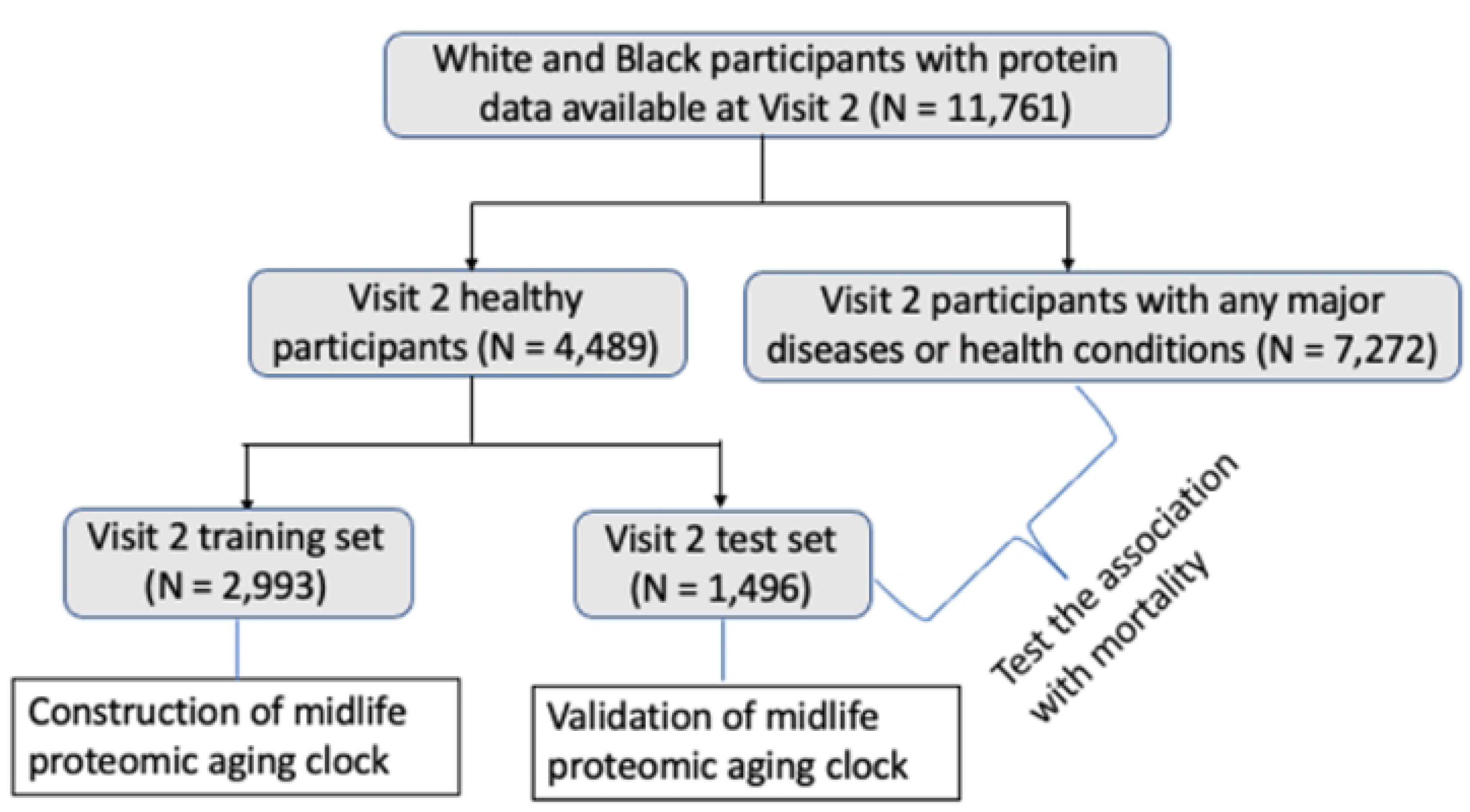
Study population at Visit 2 (midlife, 1990-1992, the chronological age of participants is 46-70 years); ARIC. The midlife ARIC PAC was constructed in a group of health participants in the training set and its asssociation with mortality was examined in all remaining pariticpants irrespective of health.

**Figure 2.**
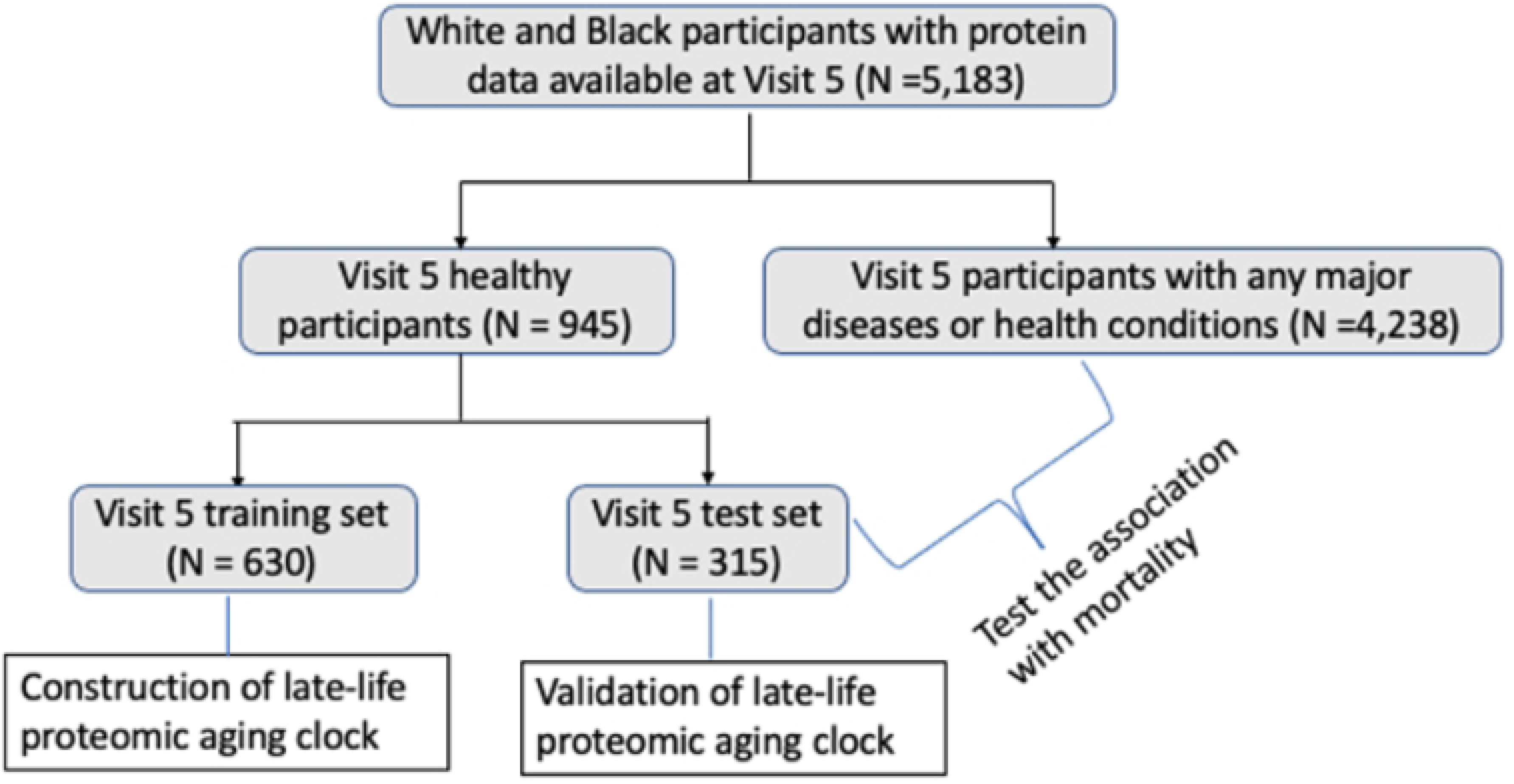
Study population at Visit 5 (late life, 2011-2013, the chronological age of participants is 66-90 years); ARIC. The late-life ARIC PAC was constructed in a group of health participants in the training set and its asssociation with mortality was examined in all remaining pariticpants irrespective of health.

### Assessment of mortality and other characteristics of interest

Deaths were ascertained through annual (semi-annual since 2012) follow-up telephone calls to participants or their proxies, surveillance of local hospitals, state records, and linkage to NDI through December 31, 2017 for participants in Mississippi or through December 31, 2019 for participants in other centers (36). All-cause mortality was defined as death resulting from any cause. CVD mortality, cancer mortality, and LRD mortality were defined based on the underlying cause of death using *International Classification of Diseases*, *Ninth Revision*, codes (ICD-9 codes) 390–459 or *International Classification of Diseases, Tenth Revision*, codes (ICD-10 codes) I00–I99 for CVD deaths, ICD-9 codes 140-239 or ICD-10 codes C00-C97 for cancer deaths, and ICD-9 codes 466 and 480-519 or ICD-10 codes J10-J98 for LRD deaths.

Other characteristics of interest included demographic, lifestyle, and medical characteristics. Namely chronological age, sex, race, study center, education, smoking status, pack-years of smoking, alcohol intake, body mass index (BMI), physical activity, aspirin use, hormone replacement therapy (HRT) in females (only at Visit 2; this variable is not available at Visit 5), and eGFR (26). Education attainment was collected at Visit 1. Physical activity was collected at Visit 1 (used as physical activity at Visit 2 in this study), and Visit 5. The other variables listed above were collected at both Visit 2 and Visit 5. Detailed procedures for assessing these characteristics are described in the **S1 Appendix**.

### Statistical analysis

#### Development of PACs

To construct ARIC PACs in midlife (Visit 2) and late life (Visit 5), we randomly selected two-thirds of healthy participants at each visit and used them as the training set at the corresponding visit. The remaining one-third of healthy participants were used as the test set (**Fig 1 and Fig 2**). We utilized the training set to train PACs against chronological age and obtain the appropriate hyperparameter values and weight for each aptamer: 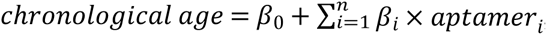, where *aptamer_i_* is the level of the *ith* aptamer. We used the test set to examine the Pearson correlation (r) between PAC and chronological age and median absolute error (MAE) to validate each PAC.

#### Construction of midlife PACs in the Visit 2 training set

Using the Visit 2 training set, we constructed the midlife ARIC PAC using elastic net regression (alpha=0.5) and with log2-transformed proteins. Lambda value was selected based on 10-fold cross-validation. We chose elastic net regression because it combines the penalties from both Lasso and Ridge regressions, and most previous aging clocks, including PACs and epigenetic clocks, were constructed using elastic net regression. Using the Visit 2 proteomics data, we also trained four other midlife PACs by applying different penalized regression methods and various protein transformations (described in **S2 Table**). For instance, one of the created PACs accounted for the potential nonlinear associations between proteins and chronological age by including both the square term and cubic term of each aptamer. Those four PACs were strongly correlated (r≥0.97) with the midlife ARIC PAC that was constructed using the simplest protein transformation (**S3 Table**). Therefore, the simplest ARIC PAC was used for further investigation.

In addition to the ARIC PAC, we also computed three published PACs in midlife: Lehallier’s (6), Tanaka’s (5), and Sathyan’s PACs (19). In our study, we computed Sathyan’s PAC using the published weights (19). For Lehallier’s and Tanaka’s PACs, we had to estimate ARIC weights specific to these PACs using Ridge regression in the training set because ARIC did not include all the aptamers reported in these PACs (**S1 Table**). The lambda value for Ridge regression was selected based on 10-fold cross-validation.

#### Construction of late-life PACs in the Visit 5 training set

Because hypertension is one of the most common conditions in older persons in the United States (37), to construct the late-life ARIC PAC, we additionally included participants with controlled hypertension as healthy participants. Controlled hypertension was defined as the measured diastolic blood pressure being below 90 and the measured systolic blood pressure being below 140 while the participant is on medication (38). Adding these participants increased the number of healthy participants by 95% (462 participants) but did not change the PAC’s performance as shown in **S4 Table**.

Using the Visit 5 training set, we constructed the late-life ARIC PAC using elastic net regression (alpha=0.5), the same approach as for the midlife ARIC PAC. Lambda value was selected based on 10-fold cross-validation. In addition to the late-life ARIC PAC, we computed the late-life Lehallier’s and Tanaka’s PACs using ARIC weights estimated using the Visit 5 training set by applying Ridge regression as discussed above and we computed the Sathyan’s PAC using the published weights.

#### Internal validation of PACs and examining associations with mortality

In the remaining 8,768 participants at Visit 2 (**Fig 1**) and 4,553 participants at Visit 5 (**Fig 2**) after excluding the training set, we computed PACs at the corresponding visits using the weighted sum of proteins determined in the training set. We internally validated each PAC in the test set of healthy participants at the corresponding visits by computing the Pearson correlation between PAC and chronological age at that visit and MAE.

In all the remaining participants at each visit, to capture the PACs’ effects independent of chronological age, we created age acceleration for each PAC as residuals after regressing PAC on chronological age (39). Demographic, lifestyle, and medical characteristics were examined across quartiles of age acceleration as mean (SD) or percentage (%). To further investigate PACs, we examined the associations between PACs and mortality. We used Cox proportional hazards regression to calculate hazard ratios (HRs) and 95% confidence intervals (CIs) for mortality from all-cause, CVD, cancer, and LRD with age acceleration. For the associations with CVD mortality, cancer mortality, and LRD mortality, deaths from other causes were treated as competing events using the Fine and Gray method (40, 41). We modeled age acceleration as a continuous variable because there was no evidence of nonlinearity observed when we applied cubic splines. For each participant, the total person-years were determined from the date of blood collection (at Visit 2 or Visit 5, depending on the analysis) until death, censoring, or the end of follow-up (either December 31, 2017 for participants from Mississippi or December 31, 2019 for participants from other centers), whichever occurred first. The proportional hazards assumption, examined by the graphical methods using log-log survival curves with age acceleration dichotomized at the median, was not violated in any regression models. The model was adjusted for chronological age, sex, joint terms for race and study center (Black participants from Mississippi; Black participants from any other centers; White participants from Maryland; White participants from North Carolina; and White participants from Minnesota), education, BMI, smoking status, pack-years of smoking, alcohol intake, physical activity, HRT (at Visit 2 only), diabetes, hypertension, and prevalent CVD, and eGFR (fully-adjusted model). These variables were associated with either age acceleration or risk of mortality. To confirm these variables as potential confounders, we computed the magnitude of R squared by regressing age acceleration for both the midlife and late-life ARIC PACs on these variables at the corresponding visits in the model adjusted for chronological age (**S6 Table**). We did not adjust for aspirin use because aspirin use had no association with midlife or late-life age acceleration for ARIC PACs and aspirin use explained <0.0015 of variance in both midlife and late-life age acceleration (**S6 Table**). In this study, we found that HRs (95% CIs) for mortality were the same in the age-adjusted and fully-adjusted models. Thus, we reported results for the fully adjusted model.

We also examined whether the change in age acceleration from midlife (Visit 2) to late life (Visit 5), computed as the age acceleration for the late-life ARIC PAC minus the age acceleration for the midlife ARIC PAC, was associated with all-cause mortality and cause-specific mortality types using Cox proportional hazard regression. For each participant, the total person-years was determined from Visit 5 date until death, censoring, or the end of follow-up. For this analysis, we additionally adjusted for midlife age acceleration. Also, we examined whether the associations with mortality were modified by midlife age acceleration (continuous variable) using a multiplicative term between the change in age acceleration and midlife age acceleration. For the change in age acceleration, we only examined the change based on the ARIC PACs because the ARIC and published PACs showed similar associations with all mortality types at each visit.

In addition to studying the associations with mortality, we examined if midlife lifestyle and medical characteristics (Visit 2) were associated with late-life age acceleration (Visit 5). This analysis was conducted using multivariable linear regression and midlife participants’ characteristics including: chronological age, sex, race, education, BMI, smoking status, pack-years of smoking, alcohol intake, physical activity (at Visit 1), HRT use, diabetes, hypertension, CVD, and eGFR were included into the model simultaneously.

Finally, we tested whether or not the exclusion of the training set influenced the associations between PACs and mortality. We examined this by comparing the associations for Sathyan’s PAC in all participants and in participants after excluding the training set. We used Sathyan’s PAC rather than other published PACs because all the proteins reported in Sathyan’s PAC were measured in ARIC and we were able to calculate Sathyan’s PAC using published weights.

#### Exploratory analyses

In an exploratory analysis, we examined whether sex, race, or chronological age (in tertiles) modified the associations of age acceleration with all-cause mortality, CVD mortality, and cancer mortality by including a multiplicative term between age acceleration and the variable of interest in the corresponding models. We did not examine LRD mortality due to the limited number of LRD deaths.

In the second exploratory analysis, we examined the association between age acceleration for the midlife ARIC PAC and the 10-year risk of death as this may be important for clinical screening. We tested the 10-year risk for midlife PAC only, because the follow-up period starting from late life was less than 10 years. Here we examined the midlife ARIC PAC only, because ARIC and published PACs showed similar associations with all mortality types.

In this study, PACs were constructed using R (version 4.1.2, package “glmnet”), and all the other analyses were performed using SAS 9.4 (RRID: SCR_008567).

## Results

### Midlife PACs

After excluding the Visit 2 training set, the remaining participants at Visit 2 (midlife) were on average 58.1±5.7 years old, 54.6% were female, and 27.1% were identified as Black.

#### Pearson correlation coefficients between PACs and chronological age in midlife

Elastic net regression selected 788 aptamers for the midlife ARIC PAC (**Table 1**). In the Visit 2 test set, the midlife ARIC PAC was correlated with chronological age (r=0.80, MAE=2.19 years, **Table 1 and Fig 3a**). Of the three midlife published PACs, Lehallier’s PAC (r=0.76, **Table 1 and Fig 3b**) had a slightly higher correlation with chronological age than Tanaka’s (r=0.66) and Sathyan’s PACs (r=0.58) (**S5 Table and S1 Fig**). The midlife ARIC PAC was strongly correlated with the midlife Lehallier’s (r=0.89), Tanaka’s (r=0.77), and Sathyan’s PACs (r=0.71) (**S3 Table**).

**Figure 3.**
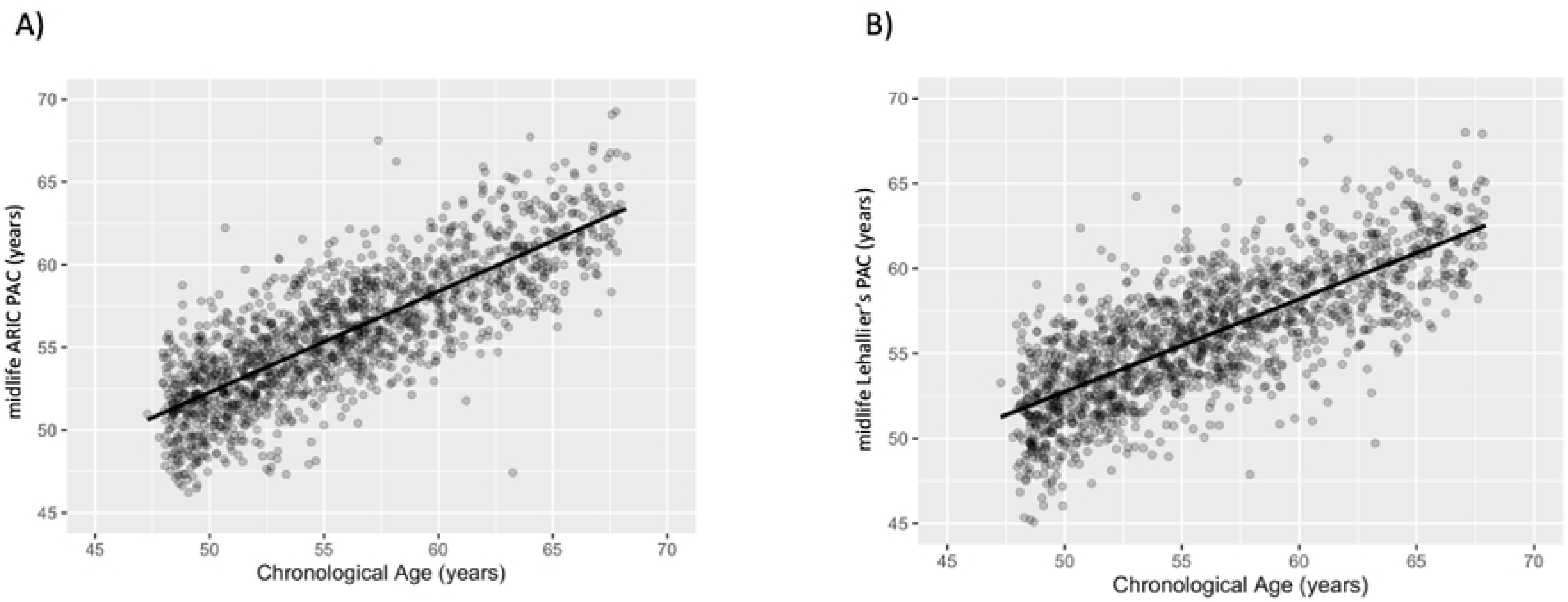
Pearson correlation (r) between the midlife ARIC and Lehallier’s PACs and chronological age in the Visit 2 test set of healthy participants. The x-axis depicts chronological age. The y-axis represents proteomic aging clock (PAC). (A) The midlife ARIC PAC was constructed using healthy participants from ARIC. The correlation between the midlife ARIC PAC and chronological age was 0.80. (B) Lehallier’s PAC was computed using ARIC weights obtained from Ridge regression based on proteins available in ARIC. The correlation between midlife Lehallier’s PAC and chronological age was 0.76.

**Table 1.** Pearson correlation between the midlife and late-life ARIC and Lehallier’s PACs and chronological age and MAE, ARIC.

| *The midlife ARIC and published PACs; Visit 2 (N = 2,993 in training set, N = 1,496 in test set)* |  |  |
| --- | --- | --- |
|  | midlife ARIC PAC | midlife Lehallier's PAC |
| Number of aptamers in PAC | 788 | 415 |
| Hyperparameter value (lambda) | 0.11 | 1.03 |
| Correlation in the training set<sup>a</sup> | 0.92 | 0.82 |
| Correlation in the test set<sup>a</sup> | 0.80 | 0.76 |
| MAE in the training set<sup>a</sup> | 1.50 | 2.13 |
| MAE in the test set<sup>a</sup> | 2.19 | 2.39 |
| *The late-life ARIC and published PACs; Visit 5 (N = 630 in training set, N = 315 in test set)* |  |  |
|  | late-life ARIC PAC | late-life Lehallier's PAC |
| Number of aptamers in PAC | 135 | 415 |
| Hyperparameter value (lambda) | 0.46 | 4.42 |
| Correlation in the training set<sup>a</sup> | 0.84 | 0.84 |
| Correlation in the test set<sup>a</sup> | 0.71 | 0.63 |
| MAE in the training set<sup>a</sup> | 1.47 | 1.71 |
| MAE in the test set<sup>a</sup> | 2.36 | 2.37 |

Abbreviations: MAE – median absolute error; PAC – proteomic ageing clock.
<sup>a</sup>Among healthy participants at Visit 2 and Visit 5, we randomly selected two-thirds of healthy participants at each visit and used them as the training set at the corresponding visit; the remaining one-third of healthy participants at each visit was used as the test set at the corresponding visit.

#### Distributions of midlife characteristics across quartiles of midlife age acceleration

Distributions of midlife characteristics (Visit 2) across quartiles of midlife age acceleration are shown in **Table 2** and **S7 Table**. Among the 8,768 participants in midlife (all Visit 2 participants after excluding the Visit 2 training set), the range of age acceleration was from −14.0 to +24.2 years for the midlife ARIC PAC. The distributions of characteristics including HRT use, prevalent CVD, and eGFR were in the same direction across age acceleration for the midlife ARIC and published PACs (**Table 2** and **S7 Table**). However, the distributions of gender, race, education, BMI, current smoking, aspirin use, prevalent hypertension, and prevalent diabetes were different across different PACs (**Table 2** and **S7 Table**). The difference may be because different PACs capture different aspects of aging.

**Table 2.**
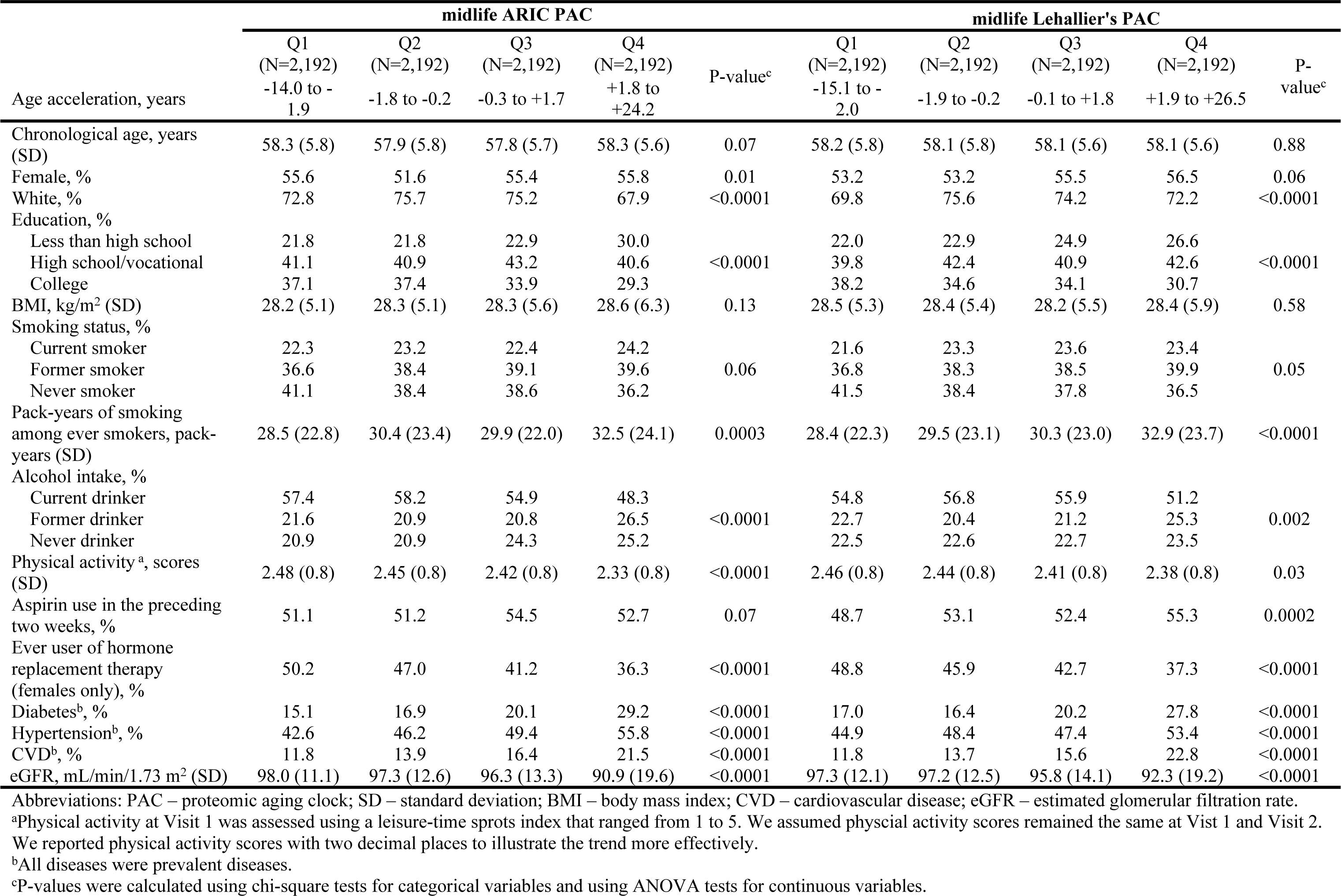
Midlife participants’ characteristics across quartiles of age acceleration for the midlife ARIC and Lehallier’s PACs; ARIC.

|  | midlife ARIC PAC |  |  |  | P-value<sup>c</sup> | midlife Lehallier's PAC |  |  |  | P-value<sup>c</sup> |
| --- | --- | --- | --- | --- | --- | --- | --- | --- | --- | --- |
|  | Q1<br>(N=2,192)<br>-14.0 to -<br>1.9 | Q2<br>(N=2,192)<br>-1.8 to -0.2 | Q3<br>(N=2,192)<br>-0.3 to +1.7 | Q4<br>(N=2,192)<br>+1.8 to<br>+24.2 |  | Q1<br>(N=2,192)<br>-15.1 to -<br>2.0 | Q2<br>(N=2,192)<br>-1.9 to -0.2 | Q3<br>(N=2,192)<br>-0.1 to +1.8 | Q4<br>(N=2,192)<br>+1.9 to +26.5 |  |
| Age acceleration, years |  |  |  |  |  |  |  |  |  |  |
| Chronological age, years (SD) | 58.3 (5.8) | 57.9 (5.8) | 57.8 (5.7) | 58.3 (5.6) | 0.07 | 58.2 (5.8) | 58.1 (5.8) | 58.1 (5.6) | 58.1 (5.6) | 0.88 |
| Female, % | 55.6 | 51.6 | 55.4 | 55.8 | 0.01 | 53.2 | 53.2 | 55.5 | 56.5 | 0.06 |
| White, % | 72.8 | 75.7 | 75.2 | 67.9 | <0.0001 | 69.8 | 75.6 | 74.2 | 72.2 | <0.0001 |
| Education, % |  |  |  |  |  |  |  |  |  |  |
| Less than high school | 21.8 | 21.8 | 22.9 | 30.0 |  | 22.0 | 22.9 | 24.9 | 26.6 |  |
| High school/vocational | 41.1 | 40.9 | 43.2 | 40.6 | <0.0001 | 39.8 | 42.4 | 40.9 | 42.6 | <0.0001 |
| College | 37.1 | 37.4 | 33.9 | 29.3 |  | 38.2 | 34.6 | 34.1 | 30.7 |  |
| BMI, kg/m<sup>2</sup> (SD) | 28.2 (5.1) | 28.3 (5.1) | 28.3 (5.6) | 28.6 (6.3) | 0.13 | 28.5 (5.3) | 28.4 (5.4) | 28.2 (5.5) | 28.4 (5.9) | 0.58 |
| Smoking status, % |  |  |  |  |  |  |  |  |  |  |
| Current smoker | 22.3 | 23.2 | 22.4 | 24.2 |  | 21.6 | 23.3 | 23.6 | 23.4 |  |
| Former smoker | 36.6 | 38.4 | 39.1 | 39.6 | 0.06 | 36.8 | 38.3 | 38.5 | 39.9 | 0.05 |
| Never smoker | 41.1 | 38.4 | 38.6 | 36.2 |  | 41.5 | 38.4 | 37.8 | 36.5 |  |
| Pack-years of smoking among ever smokers, pack-years (SD) | 28.5 (22.8) | 30.4 (23.4) | 29.9 (22.0) | 32.5 (24.1) | 0.0003 | 28.4 (22.3) | 29.5 (23.1) | 30.3 (23.0) | 32.9 (23.7) | <0.0001 |
| Alcohol intake, % |  |  |  |  |  |  |  |  |  |  |
| Current drinker | 57.4 | 58.2 | 54.9 | 48.3 |  | 54.8 | 56.8 | 55.9 | 51.2 |  |
| Former drinker | 21.6 | 20.9 | 20.8 | 26.5 | <0.0001 | 22.7 | 20.4 | 21.2 | 25.3 | 0.002 |
| Never drinker | 20.9 | 20.9 | 24.3 | 25.2 |  | 22.5 | 22.6 | 22.7 | 23.5 |  |
| Physical activity<sup>a</sup>, scores (SD) | 2.48 (0.8) | 2.45 (0.8) | 2.42 (0.8) | 2.33 (0.8) | <0.0001 | 2.46 (0.8) | 2.44 (0.8) | 2.41 (0.8) | 2.38 (0.8) | 0.03 |
| Aspirin use in the preceding two weeks, % | 51.1 | 51.2 | 54.5 | 52.7 | 0.07 | 48.7 | 53.1 | 52.4 | 55.3 | 0.0002 |
| Ever user of hormone replacement therapy (females only), % | 50.2 | 47.0 | 41.2 | 36.3 | <0.0001 | 48.8 | 45.9 | 42.7 | 37.3 | <0.0001 |
| Diabetes<sup>b</sup>, % | 15.1 | 16.9 | 20.1 | 29.2 | <0.0001 | 17.0 | 16.4 | 20.2 | 27.8 | <0.0001 |
| Hypertension<sup>b</sup>, % | 42.6 | 46.2 | 49.4 | 55.8 | <0.0001 | 44.9 | 48.4 | 47.4 | 53.4 | <0.0001 |
| CVD<sup>b</sup>, % | 11.8 | 13.9 | 16.4 | 21.5 | <0.0001 | 11.8 | 13.7 | 15.6 | 22.8 | <0.0001 |
| eGFR, mL/min/1.73 m<sup>2</sup> (SD) | 98.0 (11.1) | 97.3 (12.6) | 96.3 (13.3) | 90.9 (19.6) | <0.0001 | 97.3 (12.1) | 97.2 (12.5) | 95.8 (14.1) | 92.3 (19.2) | <0.0001 |

Abbreviations: PAC – proteomic aging clock; SD – standard deviation; BMI – body mass index; CVD – cardiovascular disease; eGFR – estimated glomerular filtration rate.
<sup>a</sup>Physical activity at Visit 1 was assessed using a leisure-time sports index that ranged from 1 to 5. We assumed physical activity scores remained the same at Visit 1 and Visit 2. We reported physical activity scores with two decimal places to illustrate the trend more effectively.
<sup>b</sup>All diseases were prevalent diseases.
<sup>c</sup>P-values were calculated using chi-square tests for categorical variables and using ANOVA tests for continuous variables.

#### Association between midlife age acceleration and mortality

Among the 8,768 participants at Visit 2, 5,294 died by 2019 with a median follow-up of 23.8 years. Age acceleration for the midlife ARIC PAC and published PACs showed associations of similar magnitude with all mortality types (**Table 3 and S8 Table**). For the midlife ARIC PAC, a one SD (SD = 2.94 years) increase in age acceleration was associated with a 38% increased risk of all-cause mortality [95% CI: 1.34-1.42], a 20% increased risk of CVD mortality [95% CI: 1.14-1.27], and a 36% increased risk of LRD mortality [95% CI: 1.22-1.51] (**Table 3**). Neither age acceleration for the midlife ARIC PAC nor published PACs was associated with cancer mortality (**Table 3**).

**Table 3.**
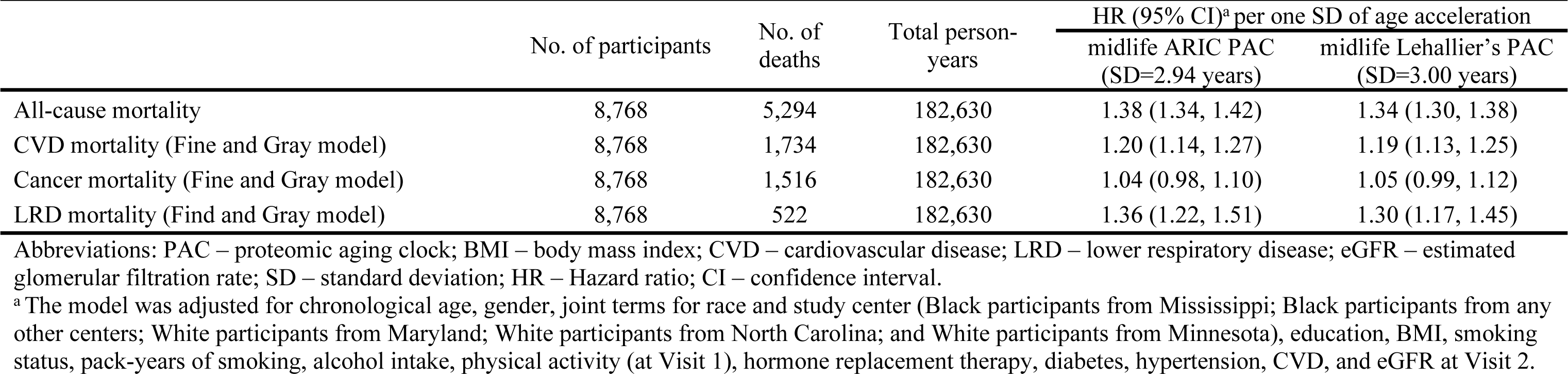
The associations between age acceleration for the midlife ARIC and Lehallier’s PACs and mortality; ARIC (1990-2019)

|  | No. of participants | No. of deaths | Total person-years | HR (95% CI)<sup>a</sup> per one SD of age acceleration |  |
| --- | --- | --- | --- | --- | --- |
|  |  |  |  | midlife ARIC PAC<br>(SD=2.94 years) | midlife Lehallier's PAC<br>(SD=3.00 years) |
| All-cause mortality | 8,768 | 5,294 | 182,630 | 1.38 (1.34, 1.42) | 1.34 (1.30, 1.38) |
| CVD mortality (Fine and Gray model) | 8,768 | 1,734 | 182,630 | 1.20 (1.14, 1.27) | 1.19 (1.13, 1.25) |
| Cancer mortality (Fine and Gray model) | 8,768 | 1,516 | 182,630 | 1.04 (0.98, 1.10) | 1.05 (0.99, 1.12) |
| LRD mortality (Find and Gray model) | 8,768 | 522 | 182,630 | 1.36 (1.22, 1.51) | 1.30 (1.17, 1.45) |

Abbreviations: PAC – proteomic aging clock; BMI – body mass index; CVD – cardiovascular disease; LRD – lower respiratory disease; eGFR – estimated glomerular filtration rate; SD – standard deviation; HR – Hazard ratio; CI – confidence interval.
<sup>a</sup> The model was adjusted for chronological age, gender, joint terms for race and study center (Black participants from Mississippi; Black participants from any other centers; White participants from Maryland; White participants from North Carolina; and White participants from Minnesota), education, BMI, smoking status, pack-years of smoking, alcohol intake, physical activity (at Visit 1), hormone replacement therapy, diabetes, hypertension, CVD, and eGFR at Visit 2.

### Late-life PACs

After excluding the Visit 5 training set, the remaining participants at Visit 5 (late life) were on average 76.5±5.3 years old, 56.3% were female, and 19.7% were identified as Black.

#### Pearson correlation coefficients between PACs and chronological age in late life

Elastic net regression selected 135 aptamers for the late-life ARIC PAC (**Table 1**). In the Visit 5 test set, the late-life ARIC PAC was correlated with chronological age (r=0.71, MAE=2.36 years, **Table 1 and Fig 4a**). The late-life Lehallier’s PAC had a correlation of 0.63 with chronological age (**Table 1 and Fig 4b**) and the late-life Tanaka’s and Sathyan’s PACs had correlations of 0.59 and 0.69 with chronological age, respectively (**S5 Table and S2 Fig**). In the Visit 5 test set, the late-life ARIC PAC was strongly correlated with the late-life Lehallier’s (r = 0.84), Tanaka’s (r = 0.79), and Sathyan’s PACs (r = 0.84) (**S9 Table**).

**Figure 4.**
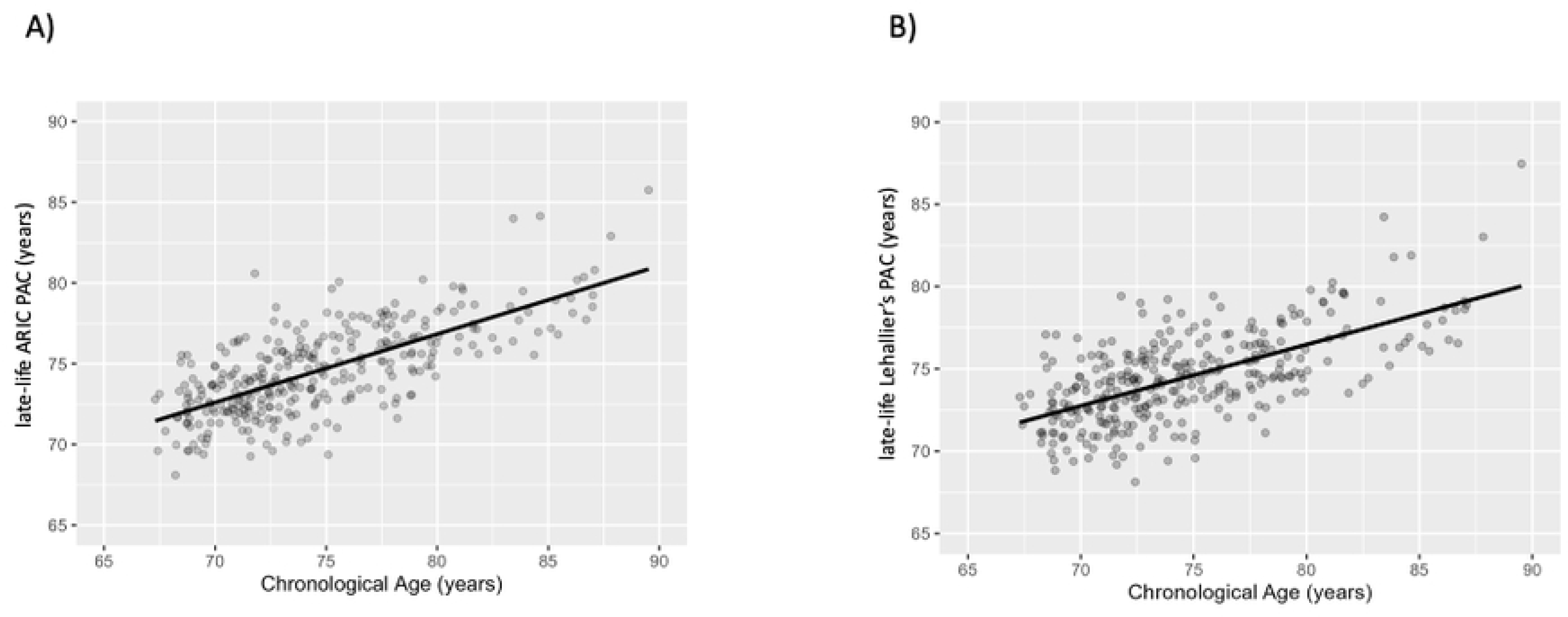
Pearson correlation (r) between the late-life ARIC and Lehallier’s PACs and chronological age in the Visit 5 test set of healthy participants, ARIC. The x-axis depicts chronological age. The y-axis represents proteomic aging clock (PAC). (A) The late-life ARIC PAC was constructed using healthy participants from ARIC. The correlation between the late-life ARIC PAC and chronological age was 0.71. (B) Lehallier’s PAC was computed using ARIC weights obtained from Ridge regression based on proteins available in ARIC. The correlation between late-life Lehallier’s PAC and chronological age was 0.63.

#### Distribution of characteristics in late life across quartiles of late-life age acceleration

Distribution of late-life characteristics (Visit 5) across quartiles of late-life age acceleration are shown in **Table 4 and S10 Table**. Among the 4,553 participants in late life (all Visit 5 participants after excluding the Visit 5 training set), the range of age acceleration was from −7.5 to +17.0 years for the late-life ARIC PAC. The distributions of characteristics including having a college-level education, physical activity, prevalent CVD, and eGFR were in the same direction across age acceleration for the late-life ARIC and published PACs (**Table 4** and **S10 Table**). However, the percentages of White participants, never smokers, and never drinkers, and the prevalences of hypertension and diabetes were different across different PACs (**Table 4** and **S10 Table**).

**Table 4.** Visit 5 participants’ characteristics across quartiles of age acceleration for late-life ARIC and Lehallier’s PACs; ARIC.

|  | late-life ARIC PAC |  |  |  |  | late-life Lehallier's PAC |  |  |  |  |
| --- | --- | --- | --- | --- | --- | --- | --- | --- | --- | --- |
|  | Q1<br>(N=1,138) | Q2<br>(N=1,138) | Q3<br>(N=1,139) | Q4<br>(N=1,138) | P-value<sup>c</sup> | Q1<br>(N=1,138) | Q2<br>(N=1,138) | Q3<br>(N=1,139) | Q4<br>(N=1,138) | P-value<sup>c</sup> |
| Age acceleration | - 7.5 to -1.8 | -1.7 to -0.2 | -0.1 to +1.4 | +1.5 to +17.0 |  | -9.1 to -1.8 | -1.7 to -0.2 | -0.1 to 1.5 | 1.6 to 14.4 |  |
| Chronological age, years (SD) | 76.9 (5.1) | 75.9 (5.0) | 76.1 (5.3) | 76.7 (5.4) | <0.0001 | 76.7 (5.2) | 76.3 (5.1) | 76.0 (5.2) | 76.8 (5.5) | 0.0009 |
| Female, % | 57.7 | 57.5 | 57.1 | 52.9 | 0.06 | 62.2 | 59.8 | 53.9 | 49.5 | <0.0001 |
| White, % | 75.9 | 83.0 | 82.5 | 79.6 | <0.0001 | 78.6 | 80.5 | 81.9 | 80.1 | 0.25 |
| Education, % |  |  |  |  |  |  |  |  |  |  |
| <High school | 13.7 | 11.7 | 15.2 | 17.0 |  | 11.3 | 13.3 | 14.8 | 18.2 |  |
| High school/vocational | 41.3 | 44.4 | 42.1 | 42.6 | 0.01 | 41.7 | 42.8 | 42.7 | 43.2 | <0.0001 |
| College | 44.9 | 43.8 | 42.7 | 40.4 |  | 47.0 | 43.9 | 42.5 | 38.6 |  |
| BMI, kg/m<sup>2</sup> (SD) | 29.1 (4.9) | 28.8 (5.2) | 28.6 (5.7) | 28.6 (6.7) | 0.15 | 28.8 (5.0) | 28.6 (5.3) | 29.2 (6.2) | 28.7 (6.1) | 0.19 |
| Smoking status, % |  |  |  |  |  |  |  |  |  |  |
| Current smoker | 4.0 | 5.1 | 6.8 | 10.0 |  | 3.6 | 6.7 | 6.4 | 9.2 |  |
| Former smoker | 54.5 | 56.8 | 50.1 | 50.8 | <0.0001 | 54.4 | 50.3 | 54.1 | 53.6 | <0.0001 |
| Never smoker | 41.4 | 38.1 | 43.2 | 39.1 |  | 42.0 | 43.0 | 39.5 | 37.2 |  |
| Pack-years of smoking among ever smokers, pack-years (SD) | 10.9 (16.3) | 11.9 (19.0) | 12.7 (21.8) | 15.4 (22.3) | <0.0001 | 10.5 (16.8) | 12.1 (21.2) | 12.2 (19.0) | 16.0 (22.2) | <0.0001 |
| Alcohol intake, % |  |  |  |  |  |  |  |  |  |  |
| Current drinker | 53.2 | 50.6 | 49.1 | 46.1 |  | 53.0 | 50.6 | 49.0 | 46.6 |  |
| Former drinker | 28.3 | 30.8 | 28.4 | 31.0 | 0.01 | 28.7 | 29.0 | 29.7 | 31.2 | 0.12 |
| Never drinker | 18.5 | 18.6 | 22.5 | 22.8 |  | 18.3 | 20.4 | 21.3 | 22.2 |  |
| Physical activity<sup>a</sup>, scores (SD) | 2.70 (0.8) | 2.64 (0.8) | 2.56 (0.8) | 2.39 (0.8) | <0.0001 | 2.71 (0.8) | 2.64 (0.8) | 2.54 (0.8) | 2.39 (0.8) | <0.0001 |
| Aspirin use in the preceding two weeks, % | 68.8 | 69.8 | 71.0 | 73.0 | 0.14 | 69.0 | 69.7 | 71.3 | 72.6 | 0.23 |
| Diabetes<sup>b</sup>, % | 40.5 | 36.3 | 34.0 | 39.2 | 0.01 | 32.4 | 35.3 | 38.5 | 43.8 | <0.0001 |
| Hypertension<sup>b</sup>, % | 75.5 | 74.9 | 75.6 | 82.1 | <0.0001 | 72.7 | 76.2 | 77.0 | 82.2 | <0.0001 |
| CVD<sup>b</sup>, % | 19.9 | 24.7 | 29.8 | 38.4 | <0.0001 | 19.6 | 23.3 | 28.4 | 41.5 | <0.0001 |
| eGFR, mL/min/1.73 m<sup>2</sup> (SD) | 77.9 (13.8) | 73.6 (14.9) | 70.6 (16.7) | 59.9 (20.2) | <0.0001 | 76.2 (14.3) | 73.6 (15.9) | 70.5 (16.8) | 61.6 (20.3) | <0.0001 |

Abbreviations: PAC – proteomic aging clock; SD – standard deviation; BMI – body mass index; CVD – cardiovascular disease; eGFR – estimated glomerular filtration rate.
<sup>a</sup>Physical activity was assessed using a leisure-time sports index that ranged from 1 to 5. We reported physical activity scores with two decimal places to illustrate the trend more effectively.
<sup>b</sup>All the diseases were prevalent diseases.
<sup>c</sup>P-values were calculated using chi-square for categorical variables and using ANOVA for continuous variables.

#### Association between late-life age acceleration and mortality

Among the 4,553 participants at Visit 5, 1,123 died by 2019 with a median follow-up of 6.53 years. Age acceleration for the late-life ARIC and three published PACs were similarly associated with all mortality types (**Table 5 and S11 Table).** For the late-life ARIC PAC, a one SD (SD=2.61 years) increase in age acceleration was associated with an increased risk of all-cause mortality [HR (95% CI) = 1.65 (1.52-1.79)], CVD mortality [HR (95% CI) = 1.37 (1.18-1.58)], cancer mortality [HR (95% CI) = 1.21 (1.02-1.44)], and LRD mortality [HR (95% CI) = 1.68 (1.32, 2.12)] (**Table 5**).

**Table 5.** The associations of age acceleration for the late-life ARIC and Lehallier’s PACs and the change in age acceleration from midlife to late life with mortality; ARIC (2011-2019)

|  | No. of participants | No. of deaths | Total person-years | HR (95%CI)<sup>a</sup> per one SD of age acceleration |  |
| --- | --- | --- | --- | --- | --- |
|  |  |  |  | late-life ARIC PAC<br>(SD = 2.61 years) | late-life Lehallier's PAC<br>(SD = 2.54 years) |
| All-cause mortality | 4,553 | 1,123 | 29,356 | 1.65 (1.52, 1.79) | 1.58 (1.46, 1.72) |
| CVD mortality (Fine and Gray model) | 4,553 | 348 | 29,356 | 1.37 (1.18, 1.58) | 1.38 (1.19, 1.62) |
| Cancer mortality (Fine and Gray model) | 4,553 | 278 | 29,356 | 1.21 (1.02, 1.44) | 1.19 (1.02, 1.40) |
| LRD mortality (Fine and Gray model) | 4,553 | 128 | 29,356 | 1.68 (1.32, 2.12) | 1.57 (1.21, 2.03) |
| **The change in age acceleration from midlife to late life<sup>b</sup>** |  |  |  |  |  |
|  | No. of participants | No. of deaths | Total person-years | HR (95%CI) per one SD of change in age acceleration<br>(SD = 2.91 years) |  |
| All-cause mortality | 2,707 | 736 | 17,081 | 1.71 (1.52, 1.94) | NA<sup>c</sup> |
| CVD mortality (Fine and Grey model) | 2,707 | 239 | 17,081 | 1.38 (1.13, 1.68) |  |
| Cancer mortality (Fine and Grey model) | 2,707 | 172 | 17,081 | 1.30 (0.98, 1.71) |  |
| LRD mortality (Fine and Gray model) | 2,707 | 94 | 17,081 | 1.46 (1.05, 2.04) |  |

Abbreviations: PAC – proteomic aging clock; SD – standard deviation; BMI – body mass index; CVD – cardiovascular disease; LRD – lower respiratory disease; eGFR – estimated glomerular filtration rate. HR – Hazard ratio; CI – confidence interval.
<sup>a</sup> The model was adjusted for chronological age, gender, joint terms for race and study center (Black participants from Mississippi; Black participants from any other centers; White participants from Maryland; White participants from North Carolina; and White participants from Minnesota), education, BMI, smoking status, pack-years of smoking, alcohol intake, physical activity, diabetes, hypertension, CVD, and eGFR at Visit 5.
<sup>b</sup> The associations for the change in age acceleration was examined among the 2,707 participants who survived until Visit 5 after excluding the training sets at Visit 2 and at Visit 5 and the model was additionally adjusted for midlife age acceleration.
<sup>c</sup> The associations between the change in age acceleration and mortality were examined using the ARIC PACs only because the ARIC PACs and published PACs showed similar associations with all mortality types.

#### Associations of the change in age acceleration from midlife to late life with mortality

The median timespan between Visit 2 and Visit 5 was 20.8 years, ranging from 18.6 to 23.5 years. Among the 2,707 participants who survived up to Visit 5 (after excluding the training sets at Visit 2 and Visit 5), the midlife and late-life ARIC PACs were correlated with each other (r=0.69) and 48.4% of participants had a greater age acceleration in late life compared to midlife. In the fully adjusted model (additionally adjusted for midlife age acceleration), the change in age acceleration from midlife to late life was associated with all-cause mortality, CVD mortality, and LRD mortality, but not cancer mortality. HRs (95% CIs) per one SD of the change in age acceleration were 1.71 (1.52-1.94) for all-cause mortality,1.38 (1.13-1.68) for CVD mortality, 1.46 (1.05-2.04) for LRD mortality, and 1.30 (0.98-1.71) for cancer mortality (**Table 5**). Midlife age acceleration did not modify the associations between the change in age acceleration and all-cause mortality (p-interaction for the multiplicative term=0.26), CVD mortality (p-interaction=0.64), LRD mortality (p-interaction=0.26), or cancer mortality (p-interaction=0.58).

#### Association between midlife participants’ characteristics and late-life age acceleration

In the multivariable analysis of midlife participants’ characteristics (Visit 2), we found that being current smokers, never drinkers, or having diabetes, hypertension, CVD, a higher BMI, higher pack-years of smoking, lower eGFR or lower physical activity in midlife were associated with higher late-life age acceleration (**Table 6**).

**Table 6.** Association^a^ between midlife participants’ characteristics and late-life age acceleration, i.e., age acceleration for the late-life ARIC PAC; ARIC.

| Midlife participants' Characteristics<sup>b</sup> | Coefficients | P-value or P-trend<sup>c</sup> |
| --- | --- | --- |
| Chronological age | -0.03 | <0.0001 |
| Male | 0.05 | 0.65 |
| Black | -0.67 | <0.0001 |
| Education |  |  |
| <High school | 0 |  |
| High school/vocational | -0.21 | 0.27 |
| College | -0.16 |  |
| BMI | 0.04 | <0.0001 |
| Smoking status |  |  |
| Never smoker | 0.00 |  |
| Former smoker | -0.25 | <0.0001 |
| Current smoker | 0.32 |  |
| Pack-years of smoking | 0.01 | 0.0001 |
| Alcohol intake |  |  |
| Never drinker | 0 |  |
| Former drinker | -0.14 | 0.04 |
| Current drinker | -0.27 |  |
| Physical activity | -0.08 | 0.13 |
| Hormone replacement therapy |  |  |
| Female never user | 0 |  |
| Female ever user | -0.23 | 0.04 |
| Male | 0 |  |
| Hypertension | 0.45 | <0.0001 |
| CVD | 0.45 | 0.006 |
| Diabetes | 1.04 | <0.0001 |
| eGFR | -0.03 | <0.0001 |

Abbreviations: PAC – proteomic aging clock; BMI – body mass index; CVD – cardiovascular disease; eGFR – estimated glomerular filtration rate.
<sup>a</sup> The association was examined among the 4,553 participants who had information on the late-life ARIC PAC (after excluding the Visit 5 training set), and participants' characteristics were included into model simultaneously.
<sup>b</sup> Physical activity at Visit 1 was assessed using a leisure-time sports index that ranged from 1 to 5. We assumed that physical activity scores remained the same at Visit 1 and Visit 2. All the other characteristics were collected at Visit 2.
<sup>c</sup> P-value for continuous variables and P-trend for categorical variables

#### Comparison of the associations between age acceleration and mortality in the full cohort and the cohort subset after excluding the training set

The magnitudes of associations of age acceleration for Sathyan’s PAC in both midlife and late life with mortality in all participants at each visit were comparable to the magnitudes of those associations in participants after excluding the training set (**S12 Table**).

### Proteins included in PACs

There are 49 common aptamers included in both the midlife and late-life ARIC PACs, accounting for 6.2% of all proteins in the midlife ARIC PAC and 36.4% of all proteins in the late-life ARIC PAC (**S3 Fig**). Four proteins were found in common across both the midlife and late-life ARIC PACs as well as the three published PACs: pleiotrophin (PTN), A disintegrin and metalloproteinase with thrombospondin motifs 5 (ADAMTS-5), macrophage metalloelastase (MMP12), and cell adhesion molecule-related/down-regulated by oncogenes (CDON).

We also identified 20 proteins in each ARIC PAC (midlife and late-life) based on the largest absolute weights of their constituting aptamers (**S13 Table)**. We found six proteins whose corresponding aptamers had the largest absolute weights in both ARIC PACs: transgelin (TAGL), WNT1-inducible-signaling pathway protein 2 (WISP-2), chordin-like protein 1 (CRDK1), collagen alpha-1(XV) chain (COF1), complement component C1q receptor (C1QR1), and pleiotrophin (PTN).

### Exploratory analyses

#### Associations between age acceleration and mortality stratified by sex, race, and chronological age

The results for the associations between midlife PACs and mortality stratified by sex, race, and chronological age are presented in **S14 Table** and **S4 Fig**. Notably, chronological age (in tertiles) statistically modified the associations of age acceleration for both the midlife ARIC and three published PACs with CVD mortality (p-interactions<0.01), and the association was strongest among participants aged 47–54 years (first tertile) (**S14 Table** and **S4 Fig**).

The results for the associations between late-life PACs and mortality stratified by sex, race, and chronological age are presented in **S15 Table** and **S5 Fig**. Sex statistically modified the association between age acceleration and cancer mortality (p-interactions≤0.04) for the late-life ARIC PAC as well as the three published PACs, and the association was stronger and significant in women for all PACs (**S15 Table** and **S5 Fig**). In addition, chronological age (in tertiles) significantly modified the association between age acceleration and CVD mortality (p-interaction=0.04) for the late-life ARIC PAC but not the published PACs (**S15 Table** and **S5 Fig**).

#### Association between age acceleration for midlife ARIC PAC and 10-year risk of death

Among the 8,768 participants in midlife, a total of 1,137 participants died within 10 years, including 430 deaths attributed to CVD, 434 to cancer, and 85 to LRD. In the fully adjusted model, a one SD (SD=2.94 years) increase in age acceleration for the midlife ARIC PAC was associated with an increased risk of all-cause mortality [HR (95% CI)=1.49 (1.41-1.58)], CVD mortality [HR (95% CI)=1.47 (1.33-1.62)], cancer mortality [HR (95% CI)=1.21 (1.09-1.34)], and LRD mortality [HR (95% CI)=1.95 (1.60-2.38)].

## Discussion

In a large prospective community-based study of White and Black individuals, the ARIC study, we tested three published PACs (5, 6, 19) and constructed and validated *de novo* PACs in midlife (46-70 years) and late life (66-90 years), using 4,955 aptamers measured by the SomaScan assay (v.4). Both the midlife and late-life ARIC PACs were developed in healthy participants and were strongly correlated with chronological age. Correlations between chronological age and the ARIC PACs were 0.80 in midlife and 0.71 in late life, which were slightly stronger compared to the correlations between chronological age and the three published PACs (Lehallier’s, Tanaka’s, and Sathyan’s) respectively (r=0.58-0.76 in midlife and r=0.59-0.69 in late life). All the HRs for the associations with mortality, including mortality from all-cause, CVD, cancer, and LRD, were very similar for the ARIC and published PACs in midlife and late life, respectively. Notably, the associations with all-cause mortality, CVD mortality, and LRD mortality were significant at each visit but stronger in late life than in midlife, and the associations with cancer mortality were significant in late life only. The change in age acceleration from midlife to late life had associations of similar magnitude with all-cause mortality and CVD mortality when compared to the associations for the late-life ARIC PAC. The HR estimate for LRD mortality was slightly lower for the change in age acceleration compared to the late-life ARIC PAC, but the confidence intervals for these two estimates largely overlapped. The change in age acceleration was not associated with cancer mortality.

In midlife we applied different penalized regressions and various transformations of proteins to develop five *de novo* ARIC PACs, including a PAC that accounted for non-linear associations between proteins and chronological age. These five PACs were highly correlated with each other. Thus, among these five PACs, we selected the midlife ARIC PAC, constructed using the simplest protein transformation, i.e., log2 transformation without any further transformation. We selected the PAC with the simplest protein transformation because, if validated, it would be easier to use this PAC in future studies. We also constructed the late-life ARIC PAC using the same method as employed for the midlife ARIC PAC. In our study, elastic net regression selected 788 aptamers for the midlife ARIC PAC and 135 aptamers for the late-life ARIC PAC. The smaller number of aptamers for the late-life ARIC PAC may be because of the smaller training set at Visit 5 (N=630) compared to the Visit 2 training set (N=2,993). With a larger training set, penalized regressions have more power to select more aptamers. This is in agreement with Sathyan’s PAC of 162 proteins, which was developed using the same SomaScan assay as in our study with a training set of 500 participants (19).

We compared associations of midlife and late-life ARIC and published PACs with mortality. Although different PACs included different proteins, the age acceleration for both ARIC and published PACs showed comparable associations with mortality at each time point. Our findings for all-cause mortality in midlife participants were similar to the findings in the InCHIANTI study (N=459, chronological age: 21-98 years) by Tanaka et al. In their study, they reported a significant association between age acceleration for Tanaka’s PAC and all-cause mortality after adjusting for chronological age, sex, and study site [HR (95% CI) per 1 SD = 1.29 (1.11-1.50)] (22). Our findings suggest that PACs consisting of different proteins may be used for predicting mortality. It will be important to understand this phenomenon in future studies.

In our study, for both ARIC and published PACs, their late-life age acceleration showed stronger associations with all mortality types than midlife age acceleration. This may be because PACs that used proteins measured in late life capture information for some biological function closer to mortality than proteins measured in midlife. It is also possible that the longer follow-up of up to 29.9 years since midlife introduced regression dilution bias (42), resulting in weaker associations with midlife PACs. Potential regression dilution bias may also explain our stronger findings in the analysis of the midlife ARIC PAC and mortality with a follow-up period restricted to 10 years compared to a total follow-up of up to 29.9 years until 2019. The association between the midlife ARIC PAC and cancer mortality became significant when participants were followed for a maximum of 10 years. The associations with all-cause mortality, CVD mortality, and LRD mortality were stronger compared to the associations observed for participants followed up to 2019. In summary, our results underscored the potential of PACs to predict both biological age and mortality in midlife and late life. Our results also suggested that PACs may be useful to predict the 10-year risk of death in a clinical context.

Our findings showed that midlife individuals who were current smokers (compared to never smokers), as well as those with higher (vs. lower) BMI, lower (vs. higher) eGFR, and age-related diseases, such as CVD, hypertension, and diabetes in midlife, were associated with higher age acceleration in late life. In addition, a larger change in age acceleration from midlife to late life was associated with an increased risk of all-cause mortality, CVD mortality, and LRD mortality. Future studies should incorporate multiple time points in applying PACs to model the change in age acceleration over time.

The strengths of this population-based observational study include its prospective design with a follow-up of more than 20 years and detailed demographic and lifestyle information. Furthermore, the ARIC cohort includes a diverse sample comprising both White and Black individuals, while previous studies of PACs either had small sample sizes or included mainly White individuals (5, 6, 19). Also, we compared multiple PACs regarding their correlation with chronological age and their associations with mortality. In addition, with the availability of proteomics data from two distinct visits, we were able to examine the association between the midlife to late-life change in age acceleration and mortality. Moreover, we adjusted for a broader range of confounders while previous studies of PACs only adjusted for demographic factors (19, 22). Our study has several possible limitations. First, the possibility of protein degradation during long-term storage cannot be excluded. However, the blood samples were frozen right after their collection and have never been thawed reducing the possibility of degradation. Further, no evidence of protein degradation across two visits in ARIC was shown by the similar precision of the assay from a split duplicate analysis at both visits (CVBA = 6% at Visit 2 and 7% at Visit 5) (16). Second, ARIC measured proteins in plasma, rather than other tissues, which limited the generalizability of our PACs to proteins from other tissues.

In conclusion, we developed de novo midlife and late-life PACs in a diverse population of White and Black individuals and showed that these PACs were associated with mortality risk. The magnitude of these associations is similar to the associations observed for previously published PACs, both in midlife and late life. Moreover, the change in age acceleration from midlife to late life showed comparable associations with mortality as the late-life PAC. Future studies are recommended to investigate the potential use of these PACs as biomarkers for biological age and risk stratification for age-related disease. If validated in external studies, these PACs may serve as surrogate endpoints in clinical trials of anti-aging interventions and inform physicians about the implementation of anti-aging lifestyle and therapeutic interventions.

## Data Availability

ARIC data can be accessed via BioLINCC (https://biolincc.nhlbi.nih.gov/studies/aric/) free of charge and without the need for the ARIC Study approval (accession number: HLB00020023a).

## Acknowledgment

The Atherosclerosis Risk in Communities study has been funded in whole or in part with Federal funds from the National Heart, Lung, and Blood Institute; National Institutes of Health; Department of Health and Human Services, under Contract nos. (75N92022D00001, 75N92022D00002, 75N92022D00003, 75N92022D00004, 75N92022D00005). SomaLogic Inc. conducted the SomaScan assays in exchange for the use of ARIC data. This work was supported in part by NIH/NHLBI grant R01 HL134320. Cancer data in ARIC are also supported by the National Cancer Institute (U01 CA164975 and NU58DP007114). Cancer incidence data have been provided by the Maryland Cancer Registry, Center for Cancer Surveillance and Control, Department of Mental Health and Hygiene, 201 W. Preston Street, Room 400, Baltimore, MD 21201. We acknowledge the State of Maryland, the Maryland Cigarette Restitution Fund, and the National Program of Cancer Registries (NPCR) of the Centers for Disease Control and Prevention (CDC) for the funds that helped support the availability of the cancer registry data. The authors thank the staff and participants of the ARIC study for their important contributions. This study was also supported by R01CA267977 and R21AG079242. Dr. Lutsey was partially supported by NIH/NHLBI K24 HL159246. Dr. Walker is supported by the National Institute on Aging’s Intramural Research Program. This study was funded, in part, by the National Institute on Aging Intramural Research Program. The content of this work is solely the responsibility of the authors and does not necessarily represent the official views of the National Institutes of Health.

## Supporting information

**S1 Appendix:** Assessment of diseases and characteristics of interests, as well as procedures for identifying healthy participants.

**S1 Fig:** Pearson correlation (r) between midlife Tanaka’s and Sathyan’s proteomic aging clocks (PACs) and chronological age in healthy participants of the Visit 2 test set, ARIC. The x-axis depicts chronological age. The y-axis represents proteomic aging clock (PAC). (A) Tanaka’s PAC was computed using ARIC weights obtained from Ridge regression based on proteins available in ARIC. The correlation between midlife Tanaka’s PAC and chronological age was 0.66. (B) Sathyan’s PAC was calculated using the published weights. The correlation between midlife Sathyan’s PAC and chronological age was 0.58.

**S2 Fig:** Pearson correlation (r) between late-life Tanaka’s and Sathyan’s proteomic aging clocks (PAC) and chronological age in healthy participants of the Visit 5 test set, ARIC. The x-axis depicts chronological age. The y-axis represents proteomic aging clock (PAC). (A) Tanaka’s PAC was computed using ARIC weights obtained from Ridge regression based on proteins available in ARIC. The correlation between late-life Tanaka’s PAC and chronological age was 0.59. (B) Sathyan’s PAC was calculated using the published weights. The correlation between late-life Sathyan’s PAC and chronological age was 0.69.

**S3 Fig:** Overlap of aptamers included in the midlife and late-life ARIC proteomic aging clocks (PACs). The gray circle shows the aptamers included in the midlife ARIC PAC and the yellow circle shows the aptamers included in the late-life ARIC PAC.

**S4 Fig:** Association between age acceleration for the midlife ARIC PAC and mortality stratified by race, gender, and chronological age (in tertiles); ARIC (1990–2019)

**S5 Fig:** Association between age acceleration for the late-life ARIC PAC and mortality stratified by race, gender, and chronological age (in tertiles); ARIC (2011–2019)

**S1 Table**: Description of the ARIC and published proteomic aging clocks (PACs)

**S2 Table**: Description of the de novo ARIC proteomic aging clocks (PACs) constructed in middle-aged healthy participants; ARIC

**S3 Table**: Pearson correlation coefficients between the de novo ARIC proteomic aging clocks (PACs) constructed in middle-aged healthy participants and midlife published PACs among the Visit 2 test set of healthy participants

**S4 Table**: Including/excluding participants with controlled hypertension^a^ for healthy participants at Visit 5 to construct proteomic aging clocks (PACs) using elastic net regression

**S5 Table:** Pearson correlation and median absolute error (MAE) between the midlife and late-life Tanaka’s and Sathyan’s proteomic aging clocks (PACs) and chronological age, ARIC

**S6 Table**: R squared after regressing age acceleration for the midlife and late-life ARIC proteomic aging clocks (PACs) on covariates at the corresponding visits

**S7 Table**: Visit 2 participants’ characteristics across quartiles of age acceleration for midlife Tanaka’s and Sathyan’s PACs; ARIC

**S8 Table**: The association between age acceleration for midlife Tanaka’s and Sathyan’s PACs and mortality; ARIC (1990-2019)

**S9 Table**: Pearson correlation coefficients between the late-life ARIC and published PACs in the Visit 5 test set of healthy participants

**S10 Table**: Visit 5 participants’ characteristics across quartiles of age acceleration for late-life Tanaka’s and Sathyan’s PACs; ARIC

**S11 Table:** The associations of age acceleration for the late-life Tanaka’s and Sathyan’s PACs with mortality; ARIC (2011-2019)

**S12 Table:** The associations of age acceleration for midlife and late-life Sathyan’s PAC with mortality in all ARIC participants

**S13 Table:** Top 20 proteins with the largest absolute weight in the midlife and late-life ARIC PACs

**S14 Table**: The association between age acceleration for the midlife ARIC and published PACs and mortality stratified by sex, race, and chronological age (in tertiles); ARIC (1990-2019)

**S15 Table**: The associations of age acceleration for the late-life ARIC and published PACs with mortality stratified by sex, race, and chronological age (in tertiles); ARIC (2011-2019)

## Notes

### Competing Interest Statement

The authors have declared no competing interest.

### Funding Statement

AP - R01CA267977 (https://www.nih.gov/) AP and SS - R21AG079242 (https://www.nih.gov/) ARIC has been funded in whole or in part with Federal funds from the National Heart, Lung, and Blood Institute National Institutes of Health Department of Health and Human Services, under Contract nos. (75N92022D00001, 75N92022D00002, 75N92022D00003, 75N92022D00004, 75N92022D00005). This work was supported in part by NIH/NHLBI grant R01 HL134320. Cancer data in ARIC are also supported by the National Cancer Institute (U01 CA164975 and NU58DP007114).

### Author Declarations

The ARIC study was approved by institutional review boards at each participating center and all study participants provided written informed consent.

